# The effect of anti-platelet agents on end organ dysfunction and mortality in community acquired pneumonia: A protocol for a systematic review and meta-analysis

**DOI:** 10.1101/2024.04.16.24305938

**Authors:** Sylvain A. Lother, Lana Tennenhouse, Rasheda Rabbani, Ahmed M. Abou-Setta, Nicole Askin, Alexis F. Turgeon, Srinivas Murthy, Brett L. Houston, Donald S. Houston, Asher A. Mendelson, Barret Rush, Emily Rimmer, John C. Marshall, Souradet Y. Shaw, Patrick R. Lawler, Yoav Keynan, Ryan Zarychanski

## Abstract

**Background:** Community acquired pneumonia (CAP) is a common cause of morbidity and mortality globally. Poor outcomes are driven by maladaptive inflammatory and thrombotic host responses. Effective therapies that modulate host responses are lacking. Anti-platelet medications modulate thrombotic and inflammatory pathways and improve long term outcomes in COVID-19 pneumonia, however, the role of anti-platelets in other etiologies of CAP remains uncertain.

**Methods:** We will conduct a systematic review and meta-analysis of randomized controlled trials (RCTs) and observational studies including adult patients hospitalized for non-COVID-19 community acquired pneumonia (CAP) investigating the effect of anti-platelets (ASA or P2Y12 inhibitors) vs. control on all-cause mortality. We will search electronic databases including MEDLINE® (Epub Ahead of Print and In-Process, In-Data-Review & Other Non-Indexed Citations), Embase, Cochrane Central Register of Controlled Trials (CENTRAL), clinical trial registries (clinicaltrials.gov, International Clinical Trials Registry Platform) and conference abstracts from inception to August 2023. Two blinded reviewers will extract data in parallel from included studies after title and abstract screening and application of eligibility criteria. We will use the Cochrane Risk of Bias tool and Newcastle Ottawa Scale to assess risk of bias and study quality from included studies. The primary meta-analysis will be conducted separately for RCTs and observational studies using Random effect inverse variance model. For observational studies, adjusted mortality estimates will be presented as hazard ratios (HR) or adjusted odds ratios (OR) whenever possible. Heterogeneity will be expressed using the *I*^2^ statistic. The evidence will be evaluated using the Grading of Recommendations, Assessment, Development, and Evaluations (GRADE) framework.

**Discussion:** The overall treatment effect and safety of anti-platelets in non-COVID-19 CAP will be summarized. The findings may be used to inform the relevance and potential study design of a future RCT evaluating anti-platelets in CAP. If anti-platelets are shown to be safe and effective, this research is expected to contribute to a new standard of treatment for CAP, and a paradigm shift towards targeting host responses in serious infections.

**Systematic Review Registration:** This protocol is reported in accordance with the guidelines produced by PRISMA-P. The protocol was registered with Open Science Framework on January 30, 2024 (DOI https://doi.org/10.17605/OSF.IO/H2G7C)

## BACKGROUND

Community acquired pneumonia (CAP) is a life-threatening lung infection and leading cause of hospitalization and mortality globally, accounting for 2.6 million annual deaths.^1,2,3^ Among hospitalized patients with CAP, 30-day overall mortality is 23%^4^ and 6-21% of patients require Intensive Care Unit (ICU) admission.^5,6^

Poor clinical outcomes in CAP are driven by maladaptive inflammatory and thrombotic host responses to infection. Respiratory pathogens activate the innate immune system, driving local and systemic inflammation and hypercoagulability through platelet activation, endothelial dysfunction, and immunothrombotic mechanisms, that persist for days to months after infection.^7,8^ Excessive inflammation and coagulation contribute to organ dysfunction due to micro and macro-vascular thrombosis.^9,10^

Vascular thrombosis risk is elevated in CAP and occurs in approximately 11% of patients at 30-days.^11–13^ An increased risk of venous thromboembolism (VTE) was seen in COVID-19,^14,15^ and the incidence of symptomatic VTE in COVID-19 and CAP is comparable (2.0% vs. 3.6%, respectively), and higher in mechanically ventilated patients.^16^ Influenza may be associated with an even greater risk of arterial thrombosis compared with COVID-19 (7.5% vs. 4.4%, respectively).^11^ Cardiovascular events, often driven by vascular thrombosis, complicate CAP in up to a third of hospitalizations and these events are associated with a greater than 3-fold increase in mortality.^4,13,17^

Effective therapies that modulate the host response are lacking. Anti-platelet agents including acetylsalicylic acid (ASA) and P2Y12 inhibitors such as ticagrelor, clopidogrel, and prasugrel are familiar and widely accessible medications that have pleotropic effects with anti-thrombotic and anti-inflammatory activity. These mechanisms hold promise in blunting host immunothrombotic responses to infection.

In patients hospitalized with COVID-19 pneumonia, ASA demonstrated mixed effects in clinical trials, some showing benefit and others showing no significant effect.^18–20^ Further analyses suggests that ASA may benefit individuals at longer follow up duration (180-day).^21^ In non-COVID-19 pneumonia, a small randomized control trial (RCT) showed ASA reduced myocardial infarction (MI) and cardiovascular death in hospitalized patients.^22^ Several other retrospective and prospective cohort studies have suggested a potential benefit with anti-platelet agents, however due to the heterogeneity of CAP, mixed results, and differing populations, the overall effect of anti-platelet agents in hospitalized patients with CAP remains uncertain.^23–25^

The purpose of this systematic review and meta-analysis is to evaluate the effect of anti-platelet agents on mortality, end organ failures, cardiovascular events, and bleeding for patients hospitalized with non-COVID-19 CAP. This protocol is reported in accordance with the guidelines produced by the Preferred Reporting items for Systematic Reviews and Meta-analysis for developing review protocols (PRIMSA-P).^26^

## Methods/Design

We will conduct a systematic review using methodological approaches outlined in the *Cochrane Handbook for Systematic Reviewers* and report the findings in accordance with the Preferred Reporting items for Systematic Reviews and Meta-analysis (PRISMA) criteria for RCTs, and the Meta-analysis of Observational Studies in Epidemiology (MOOSE) criteria for observational studies.^27–29^ The review question and methods have been constructed with experts in infectious diseases (AK, SL, SM, YK), critical care (AK, AM, AT, BR, JM, PL, SL, SM, RZ), hematology and thrombosis (BH, DH, EM, RZ), cardiology (PL), community health (SS), and knowledge synthesis methodology (AMAS, AT, JM, NA, RR, PL, SM, SS, RZ, YK). The roles of each team member are summarized in **Appendix 1**.

### Eligibility criteria

The PICO statement and eligibility criteria for studies to be included or excluded from the systematic review and meta-analysis are provided in **Table 1** and **Table 2**.

**Table 1:** PICO statement.

|  |  |
| --- | --- |
| P - patients | Adults aged $\geq 18$ years admitted to hospital for non-COVID-19 CAP |
| I - interventions | Patients exposed to an anti-platelet agent (ASA or P2Y12 inhibitors) |
| C - comparators | Patients not exposed to an anti-platelet agent |
| O - outcomes | Mortality, end-organ failures, and thromboembolic complications |

CAP, community acquired pneumonia

**Table 2:** Eligibility criteria for studies to be included or excluded from the systematic review and meta-analysis.

|  | Inclusion criteria | Exclusion criteria |
| --- | --- | --- |
| Study design | RCTs (including placebo controlled and open label) and observational studies (including retrospective, prospective, and bi-directional cohort studies) | Animal studies, case-control studies, cross-over studies, cluster randomized trials |
| Participants | Comparative studies of adult patients admitted to a hospital for non-COVID-19 CAP (author defined), and studies of patients with bacteremia, sepsis, acute lung injury, and/or acute respiratory distress syndrome that report comparative effectiveness data for at least one clinical outcome in the CAP subgroup (derived CAP population with exposure to an APT vs. not) | Studies who enrol $\geq 20\%$ of patients with URTI without evidence of LRTI, $\geq 20\%$ with hospital acquired pneumonia (author defined), or $\geq 20\%$ of CAP caused by COVID-19 |
| Interventions | Studies where participants received ASA or a P2Y12 inhibitor, at any dose or frequency of administration, and regardless of whether the APT was received prior to hospitalization | Studies where $\geq 20\%$ of patients are receiving DAPT, $\geq 20\%$ of patients are receiving an APT in conjunction with therapeutic-dose anticoagulation |
| Comparator | Patients not receiving an APT (placebo, standard of care, no intervention, or another non-APT intervention) | Active comparator studies (one APT compared to another APT) |
| Outcomes | Primary outcome:<br>All-cause mortality (longest follow up)<br>Secondary outcome:<br>ICU admission<br>Hospital LOS (days)<br>Use of IMV<br>Use of cardiovascular organ support (vasopressors and/or inotropes and/or ECLS)<br>Use of any organ support (composite including use of HFNO and/or NIPPV and/or IMV and/or vasopressors and/or inotropes and/or ECLS)<br>Arterial thromboembolic event (myocardial infarction and/or stroke)<br>Venous thromboembolic event (deep venous thrombosis and/or pulmonary embolism)<br>Major bleeding | None of the listed outcomes are available from the study report(s) or through communication with the study authors |
| Setting | All hospital settings where patients with CAP are cared for | Patients cared for in personal care homes, long term care, or other non-hospital settings |

APT = anti-platelet; ASA = acetyl salicylic acid; CAP = community acquired pneumonia; DAPT = dual anti-platelet; ECLS = extracorporeal life support; HFNO = high flow nasal oxygen; IMV = invasive mechanical ventilation; LOS = length of stay; LRTI = lower respiratory tract infection; NIPPV = non-invasive positive pressure ventilation; RCT = randomized controlled trial; URTI = upper respiratory tract infection

### Study design and setting

We will include parallel design RCTs (including placebo controlled and open label trials) and observational studies (including retrospective, prospective, and bi-directional study designs), presenting comparative data on the exposure and the non-exposure to the intervention in a population of patients hospitalized for non-COVID-19 CAP. The required study setting will be in hospital. Studies that include patients cared for in personal care homes, long term care, or other non-hospital settings will be excluded.

### Participants

We will include comparative studies of hospitalized adult patients (>80% of the study population is ≥ 18 years old) admitted to hospital for non-COVID-19 CAP. The diagnosis of CAP will be author defined and may include clinical or administrative definitions. Studies of patients with bacteremia, sepsis, acute lung injury, and/or acute respiratory distress syndrome that report on comparative effectiveness data for at least one clinical outcome in the CAP subgroup (derived CAP population with exposure to an anti-platelet vs. not) will also be included.

### Interventions and comparisons

Studies where patients received an anti-platelet agent (ASA or P2Y12 inhibitors, at any dose or frequency of administration) after study enrollment, regardless of whether the anti-platelet agent was received prior to hospitalization will be included. These patients will comprise the intervention group. Patients not receiving anti-platelet agents (may be receiving placebo, standard of care, no intervention, or another non-antiplatelet (non-ASA or non-P2Y12 inhibitor) intervention) will comprise the comparator group.

### Outcomes

Each outcome for RCTs and observational studies will be reported separately (not pooled) due to the inherent differences in study designs, and outcomes will be reported at longest available follow-up. The primary outcome will be all-cause mortality. For observational studies, the adjusted effect estimate on mortality will be reported as the primary analysis whenever possible, to control for possible confounding. Secondary outcome measures will include arterial thromboembolic events (myocardial infarction and/or stroke), venous thromboembolic events (deep venous thrombosis and/or pulmonary embolism), ICU admission, hospital length of stay, use of invasive mechanical ventilation (IMV), use of cardiovascular organ support (vasopressors and/or inotropes and/or extracorporeal life support), use of any organ support (composite outcome including use of non-invasive positive pressure ventilation (NIPPV) and/or IMV and/or vasopressors and/or inotropes and/or extracorporeal life support). NIV includes the use of high flow nasal oxygen (NFNO) defined as flow rate of > 30 L/min, continuous positive airway pressure (CPAP), or bi-level positive airway pressure (BiPAP). The primary safety outcome will be major bleeding (author defined).

### Search Strategy and Identification of Studies

Using the OVID platform, we will search MEDLINE® (including Epub Ahead of Print and In-Process, In-Data-Review & Other Non-Indexed Citations), Embase and the Cochrane Central Register of Controlled Trials (CENTRAL). Using individualized systematic search strategies for each database, we will identify relevant citations of studies from inception to August 2023. Search strategies will utilize a combination of controlled vocabulary (e.g.: “pneumonia”, “pneumonia, bacterial”, “pneumonia, viral”, “aspirin”) and keywords (e.g.: “pneumoni*”, “antiplatelet*”, “acetylsalicylic acid”). We will apply a modified version of the SIGN RCT and observational study filters.^30^ We will not use language restrictions for any searches. The Medline search strategy is presented in **Appendix 2**. Reference lists of relevant narrative reviews, systematic reviews, and the included studies will be searched for additional citations. To identify planned, on-going, or recently completed but unpublished clinical trials, we will search clinicaltrials.gov and the International Clinical Trials Registry Platform (ICTRP), and abstracts from relevant conferences. We will perform reference management in EndNote™ (Version 20.5, Thomson Reuters, Philadelphia, PA, USA).

### Study Selection and Data Extraction

We will examine all citations produced from the search strategy and remove duplicate citations. We will screen all title and abstracts and select studies applying the eligibility criteria, recording all decisions using the Rayyan artificial intelligence web-based platform.^31^ Full texts will be obtained for all citations that meet eligibility criteria, and these studies will be examined applying the inclusion and exclusion criteria, and the report of clinical outcomes. If full texts are not in the English language, we will translate with the online translator Google Translate (*United States, Google LLC*, 2016).^32,33^ The included studies will then be reviewed for data extraction using standardized and piloted screening and data extractions forms in Microsoft® Excel for Mac (version 16.69). Citations that cannot be excluded based on population, intervention, comparator, or study design will be moved to full text screening. Two reviewers will independently perform citation screening and full text screening, study selection, and data extraction in parallel. Conflicts between reviewers will be resolved by consensus or resolution of disagreements by a third reviewer as required.

From each study, we will extract study demographics including the study name, first author’s name, study design, publication year, journal, publication type (full article or abstract), publication language, sources of funding (industry vs. non-industry), study locations (continent), and number of centers. We will record participant characteristics in respective intervention and control arms, including total sample size, mean or median age and sex, and proportion of patients with immunocompromising condition(s), chronic respiratory or cardiovascular conditions, and history of MI or coronary artery disease (author defined). Participant diagnoses will be recorded including pathogen type (proportion bacterial, viral, or unknown), and illness severity (proportion in ICU at enrolment, baseline Sequential Organ Failure Assessment (SOFA) and/or Acute Physiology and Chronic Health Evaluation-II (APACHE-II) and/or Pneumonia Severity Index (PSI) and/or Confusion, Urea, Respiratory Rate, Blood Pressure-65 (CURB-65) score). We will record intervention characteristics, including the type of anti-platelet agent, total daily dose, proportion exposed to the anti-platelet agent prior to enrolment, whether the anti-platelet agent was started prospectively at the time of study enrolment, the duration of anti-platelet agent use after enrolment, and the proportion on concurrent therapeutic-dose anticoagulation. Lastly, we will record the duration of participant follow up and the outcomes of interest. For missing data, we will email the corresponding author for studies that are < 10 years old. If no response is received within 1 months, a second contact will be made, after which the data will considered missing.

### Risk of bias assessment

We will assess the internal validity at both the study and outcome level of included RCTs using the Cochrane Collaboration Risk of Bias tool (version 2).^34^ This tool consists of 5 domains (bias arising from the randomization process, bias due to deviations from intended interventions, bias due to missing outcome data, bias in measurement of the outcome, and bias in selection of the reported result), and a categorization of the overall risk of bias. Each separate domain will be rated “low risk”, “some concerns”, or “high risk”. The overall judgment will be low risk if all domains are judged to be low risk. The study will be judged as having some concerns, if at least one domain is judged to raise some concerns (but not at high risk of bias for any domain). Finally, the study will be judged as high risk if any one domain is judged high risk, or if the study is judged to have some concerns for multiple domains such that it substantially lowers the confidence in the results.

For observational studies, we will assess study quality using the Newcastle-Ottawa Scale (NOS).^35^ This tool consists of assessing quality within a list of numbered items in 3 domains, based on the selection of the study groups (4 numbered items), the comparability of the groups (1 numbered item), and the ascertainment of the outcome of interest (3 numbered items). High quality choices within each numbered item will be awarded a “star” (maximum 1 star per numbered item in the selection and outcome domains, maximum 2 stars for the numbered item in the comparability domain). Each study will be assigned a score of 0-9 stars. Studies scoring ≥ 7 will be considered high quality studies, 4-6 moderate quality, and ≤ 3 low quality.

Risk of bias assessment will be performed independently by two reviewers. Discrepancies between the two reviewers will be resolved by consensus or by resolution of conflicts by a third reviewer as required. Information regarding risk of bias will be used to guide sensitivity analyses and explore sources of heterogeneity.

### Data analysis

Meta-analysis will be conducted using Random effect inverse variance model. In the primary meta-analysis of the primary outcome, study level adjusted mortality estimates from observational studies will be presented as hazard ratios (HR) and adjusted odds ratios (OR) and will be pooled separately. Study level summary effect comparisons from RCTs will be presented as risk ratios (RR) with 95% confidence intervals (CI) and pooled separately. A secondary meta-analysis of the primary outcome will be performed using study level unadjusted reported effect estimates and presented as RR with 95% CI. Reported OR will be converted back to RR before conducting meta-analysis. Summary effect-estimates for secondary outcomes will be expressed as RR with 95% CI for dichotomous data and weighted mean difference (WMD) with 95% CI for continuous data. Provided there are sufficient included studies (≥ 10) in the analysis of the primary outcome, a funnel plot will be used to investigate publication bias with the Egger test and visual inspection to assess plot asymmetry.^36,37^ All analyses will be conducted using the general meta and metafor package^38^ in RStudio version 2023.09.1+494, R version 4.3.2 (R Project for Statistical Computing).

### Assessment of Heterogeneity and Subgroup Analysis

The presence of statistical heterogeneity will be expressed using the *I*^2^ statistic.^39^ In case of significant heterogeneity among studies (*I*^2^ > 50%), we will perform pre-defined subgroup analyses for the primary outcome, dependent on the number of included studies and the availability of appropriate outcomes and co-variates. These will include subgroups from studies with potential differences in methodologic considerations, included study populations, pneumonia type, or disease severity, factors that relate to the intervention, or duration of follow up (**Table 3**).

**Table 3:** Pre-defined sub-group analyses.

|  |  |
| --- | --- |
| *Methodological subgroups* | • Cochrane risk of bias for RCTs (high vs. low)<br>• Newcastle Ottawa scale study quality for observational studies (high vs. intermediate/low)<br>• Sample size (large ($n > 1,500$) vs. small ($n \leq 1500$))<br>• Data source for pneumonia diagnosis (clinical vs. administrative data) |
| *Clinical subgroups (based on population factors)* | • Study population (entire study population with CAP vs. data generated in a subgroup of patients with CAP)<br>• Etiology of CAP (bacterial vs. viral vs. unknown/mixed)<br>• Illness severity (admitted to ICU at time of study enrollment vs. not in ICU) |
| *Interventional subgroups* | • Type of anti-platelet agent used (ASA vs. other anti-platelet agent)<br>• Dose of ASA (high vs. low dose $\leq 81$ mg/d)<br>• Pre-hospital exposure to anti-platelet agent (chronic vs. new use of anti-platelet agent)<br>• Duration of anti-platelet agent use ($\leq 7$ days vs. $> 7$ days) |
| *Outcome subgroups* | • Duration of follow up for the primary outcome ($\leq 30$ days vs. $> 30$ days) |

#### External validity

We will apply the Grading of Recommendations, Assessment, Development, and Evaluations (GRADE) framework to summarize the strength of the evidence generated and make clinical practice recommendations.^40^

## DISCUSSION

The findings generated in this systematic review and meta-analysis will be summarized and presented at local and national conferences concurrently with publication in a peer reviewed journal. The findings will be used to inform the relevance and potential study design for a future large, international RCT of anti-platelet agents in CAP. Planned future studies will be presented to the Canadian Critical Care Trials Group and other national organizations that include general internists (Canadian Society of Internal Medicine), infectious disease experts (Association of Medical Microbiology and Infectious Disease Canada), and critical care experts (Critical Care Canada Forum) to seek future collaborations.

## DECLARATIONS

### Ethics approval and consent to participate

Not applicable

### Consent for publication

Not applicable

## Availability of data and materials

Search strings will be published in the included appendix. Data that is generated from extraction of included studies will be published in a peer reviewed journal and raw data will be publicly available on Open Science Forum or upon request with the corresponding author.

## Author contributions

Each author contributed to creating, editing, and approving the final draft of this manuscript. Individual roles of each co-author is detailed in Appendix 1.

## Data Availability

There is no data presented in this manuscript as it is a protocol for a systematic review and meta-analysis

## Acknowledgements

Not applicable

## APPENDIX

### Appendix 1 Systematic review team members

The review will be coordinated by a clinician scientist with infectious diseases and critical care training (SL), including development of the review question, literature search strategy, screening of relevant studies, data extraction and analysis, and preparation of the final manuscript. A second blinded reviewer (LT) with internal medicine training will screen relevant studies, extract data, and analyze risk of biases in duplicate. Experts from a variety of fields will provide content expertise, including infectious diseases (YK, AK, SM), critical care (AK, PL, AM, BR, SM, JM, AT, RZ), hematology and thrombosis (BH, RZ, EM, DH), cardiology (PL), community health and epidemiology (SS), and knowledge synthesis methodology (AMAS, NA, RR, YK, PL, SS, SM, JM, AT, RZ). An experienced librarian and medical information specialist with experience in systematic review search methodology will develop and test search strategies through an iterative process in consultation with the review team (NA). Another information specialist will peer review the search strategies prior to execution using the PRESS Checklist.^41^ A senior statistician with specific expertise in meta-analysis will oversee the analysis methods (RR). A clinician scientist with systematic review expertise in clinical trials and prospective observational studies (AMAS) will provide methodological advice. Two clinician scientists with expertise in infectious diseases, hematology and critical care will provide project oversight, methodological advice, along with content expertise, and resolution of disagreements among reviewers (YK, RZ).

### Appendix 2 Search strategy for Medline (Ovid)

Ovid MEDLINE(R) and Epub Ahead of Print, In-Process, In-Data-Review & Other Non-Indexed Citations and Daily <1946 to August 22, 2023>

## PRISMA-P 2015 Checklist

This checklist has been adapted for use with systematic review protocol submissions to BioMed Central journals from Table 3 in Moher D et al: Preferred reporting items for systematic review and meta-analysis protocols (PRISMA-P) 2015 statement. *Systematic Reviews* 2015 4:1

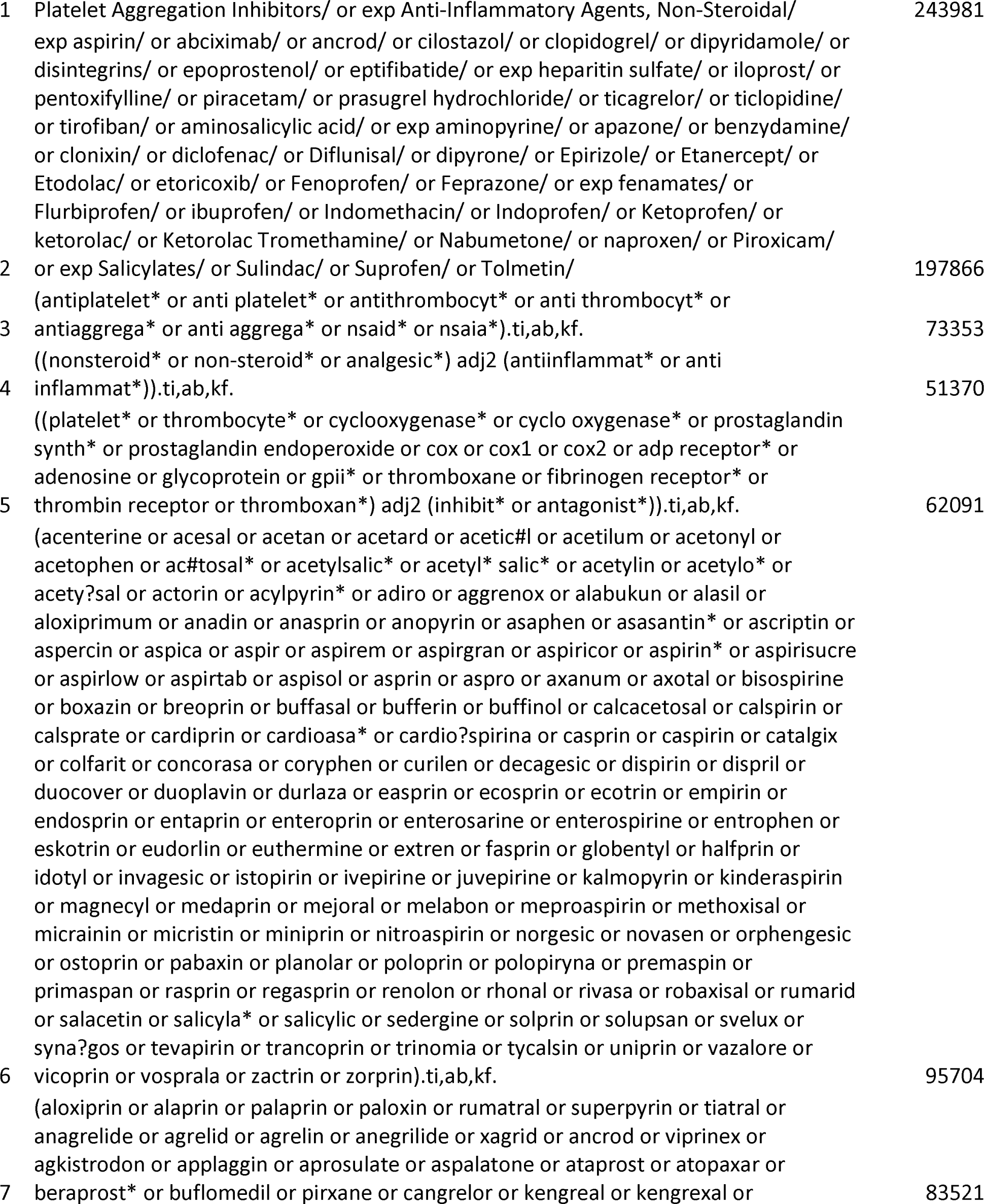
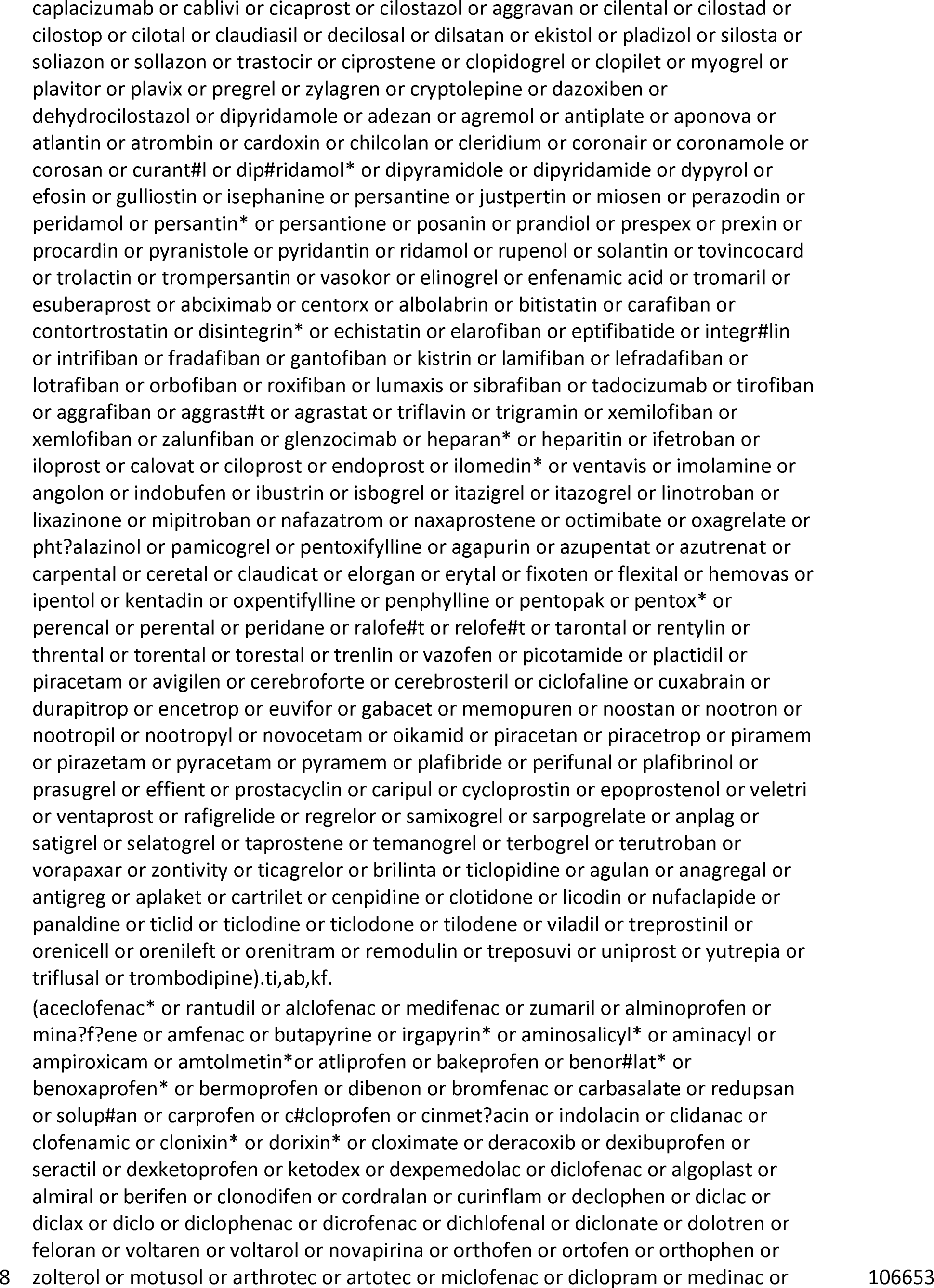
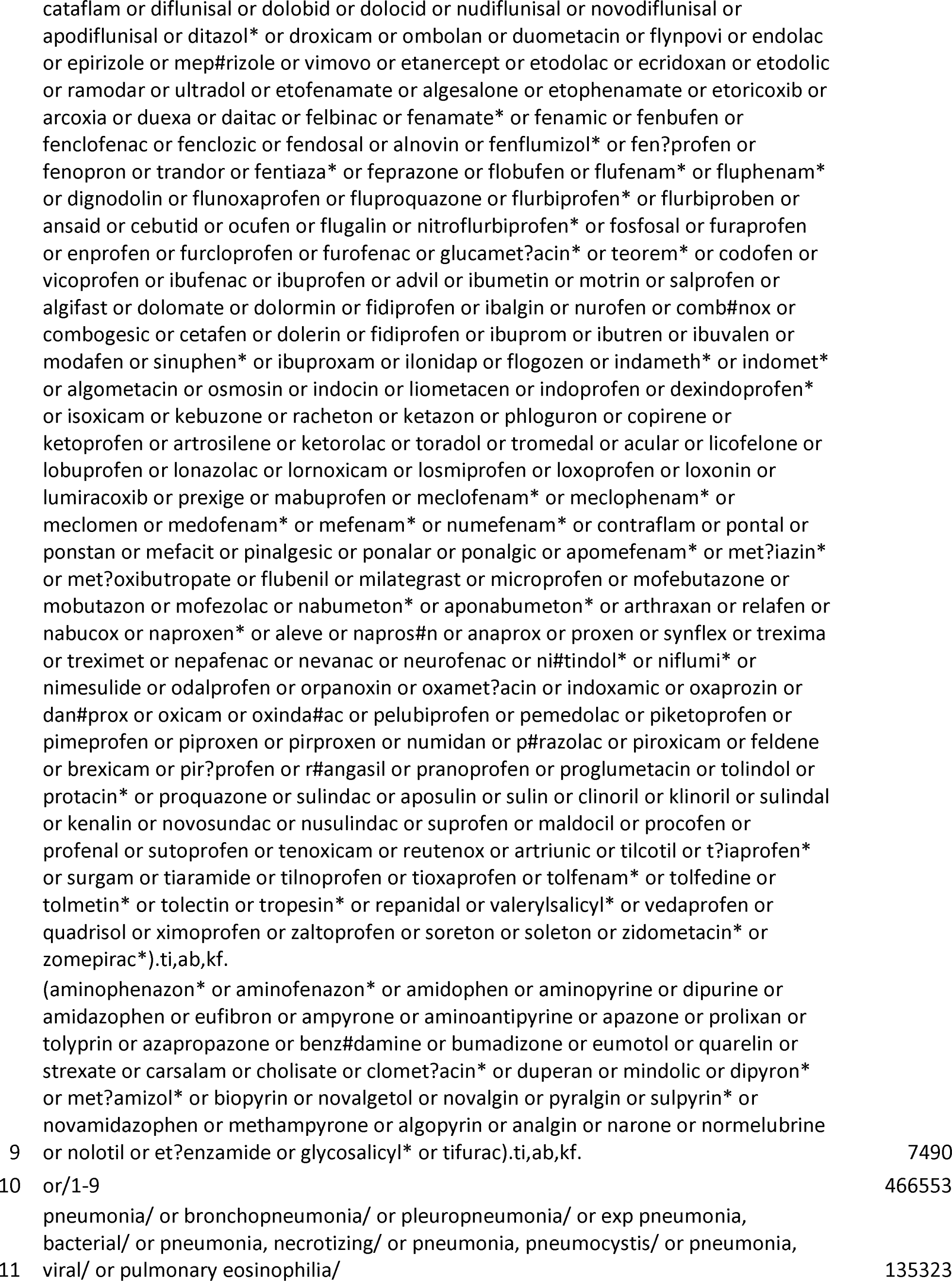
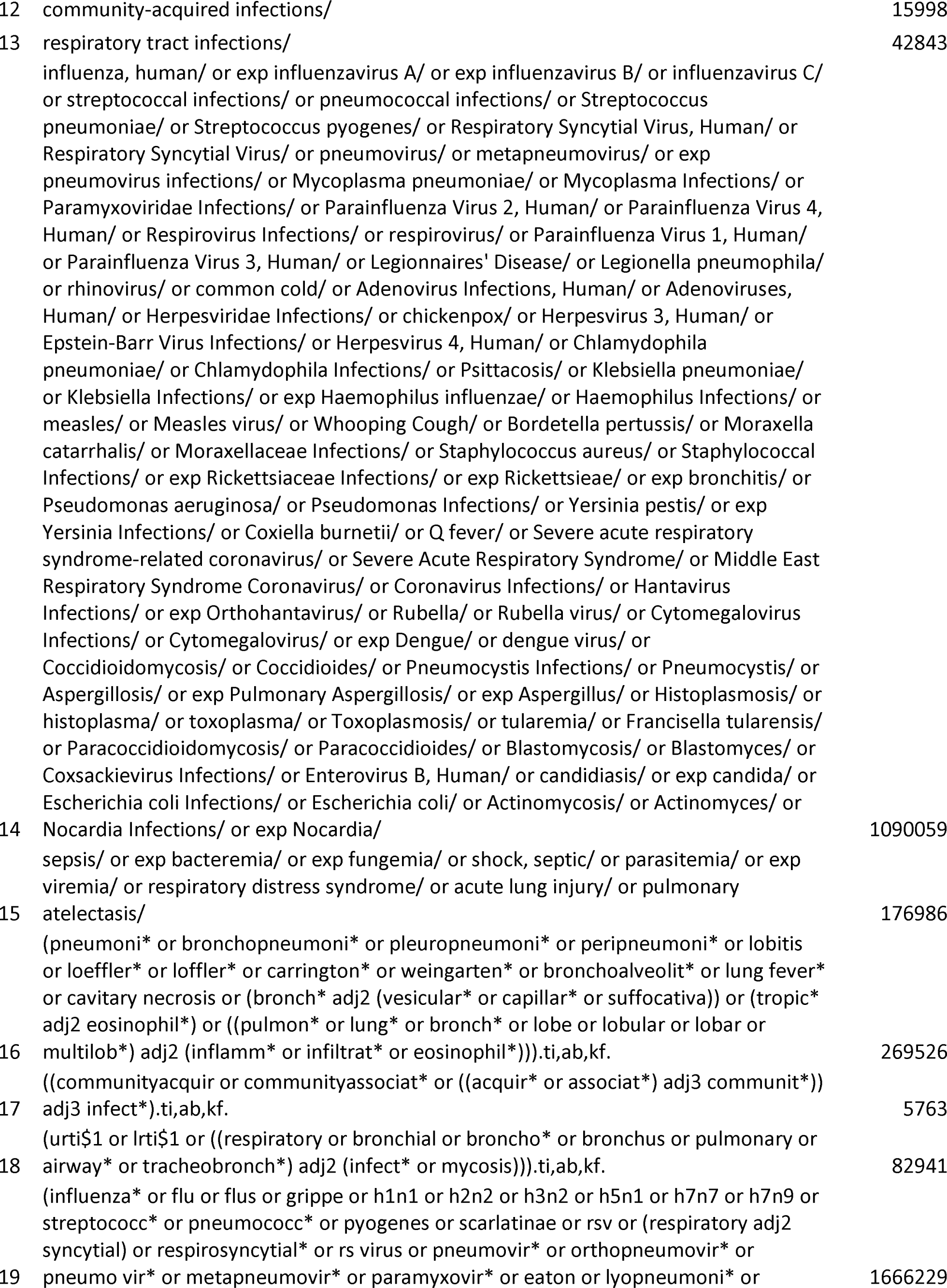
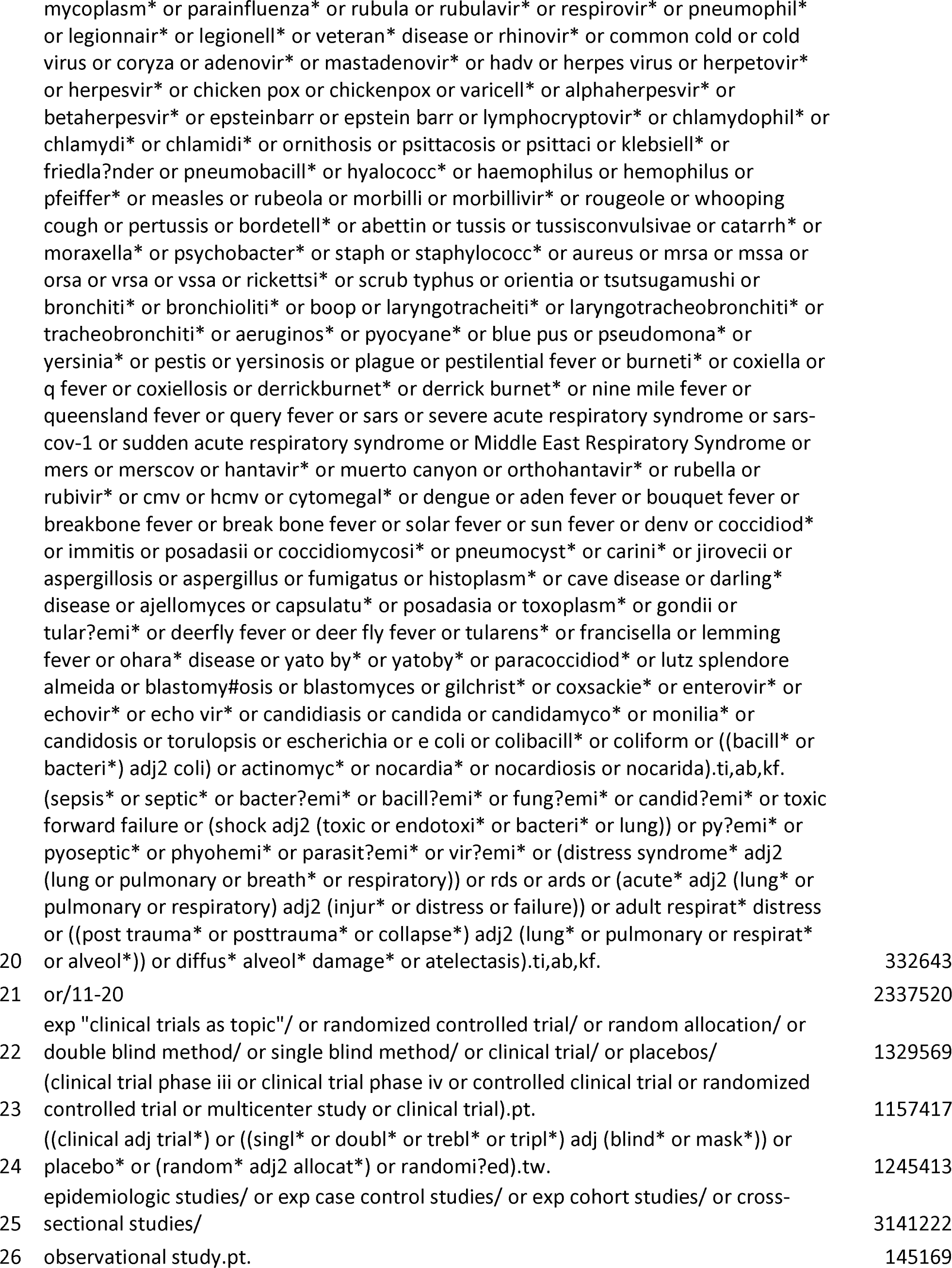
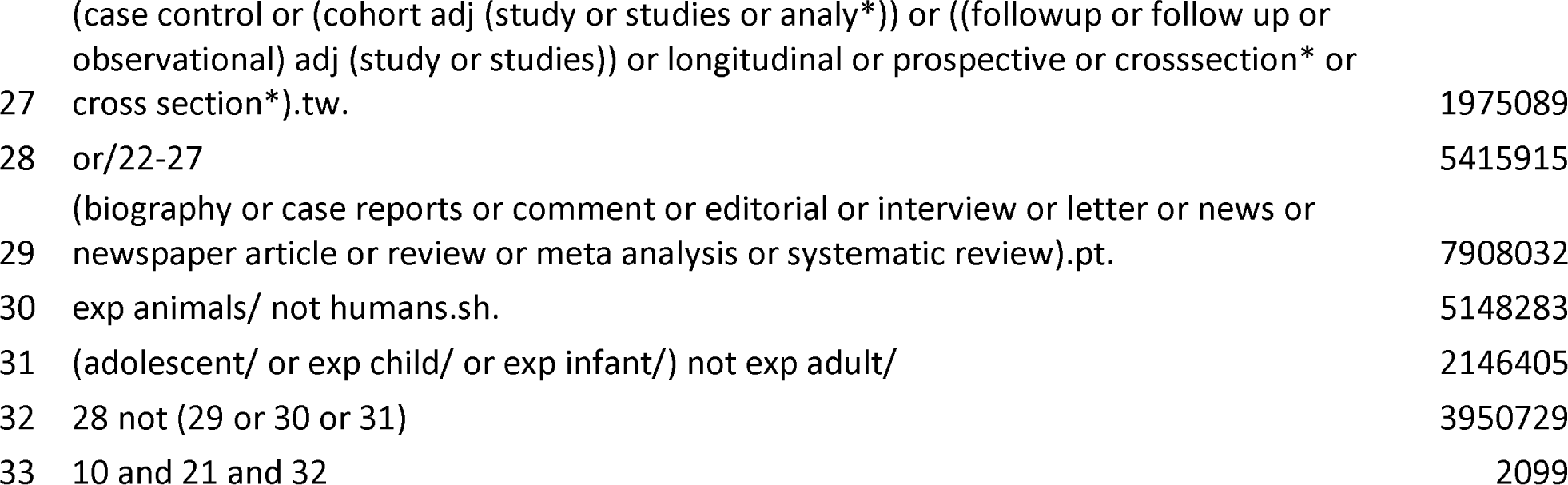

**An Editorial from the Editors-in-Chief of *Systematic Reviews* details why this checklist was adapted** - Moher D, Stewart L & Shekelle P: Implementing PRISMA-P: recommendations for prospective authors. Systematic Reviews 2016 5:15

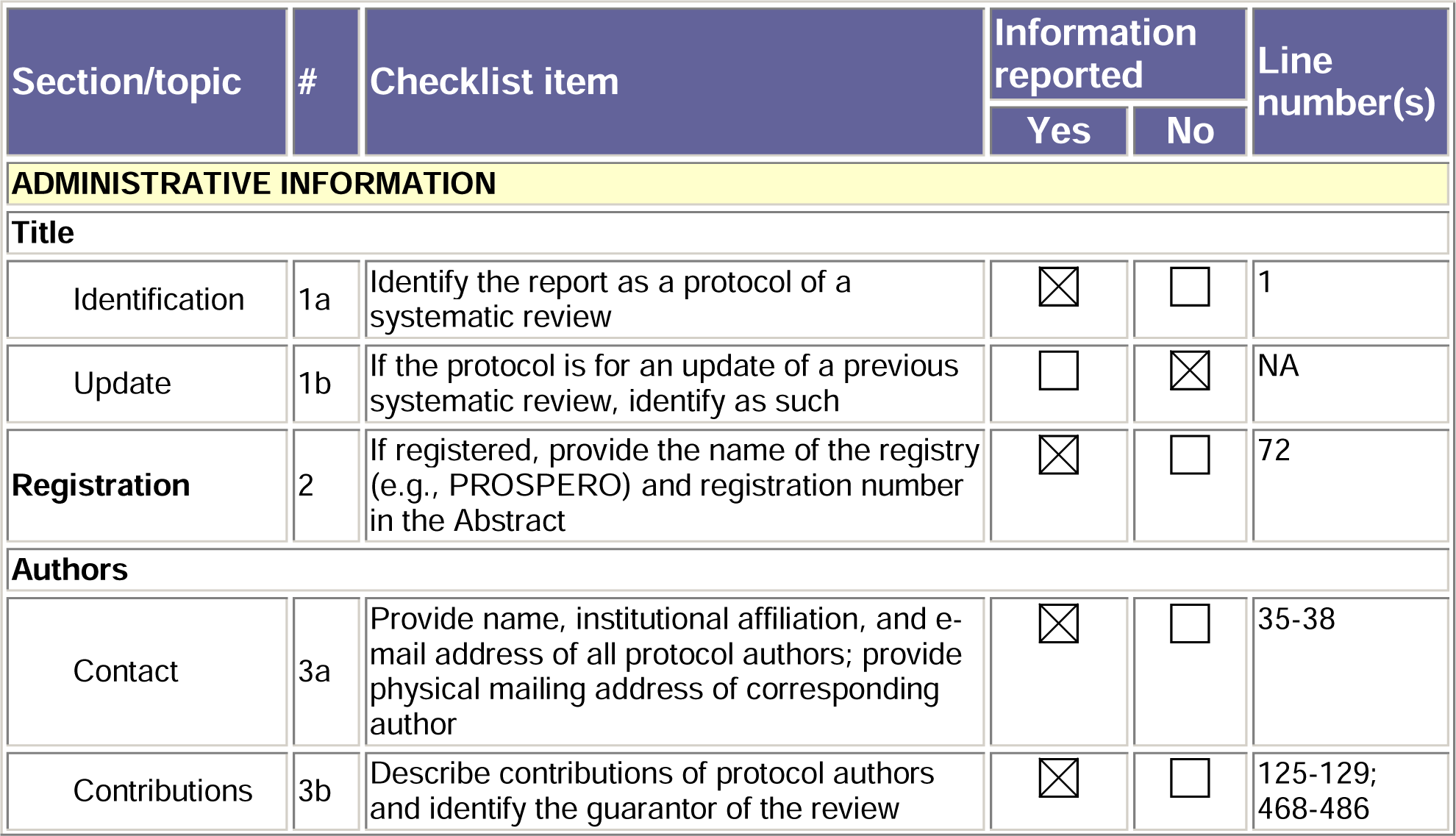
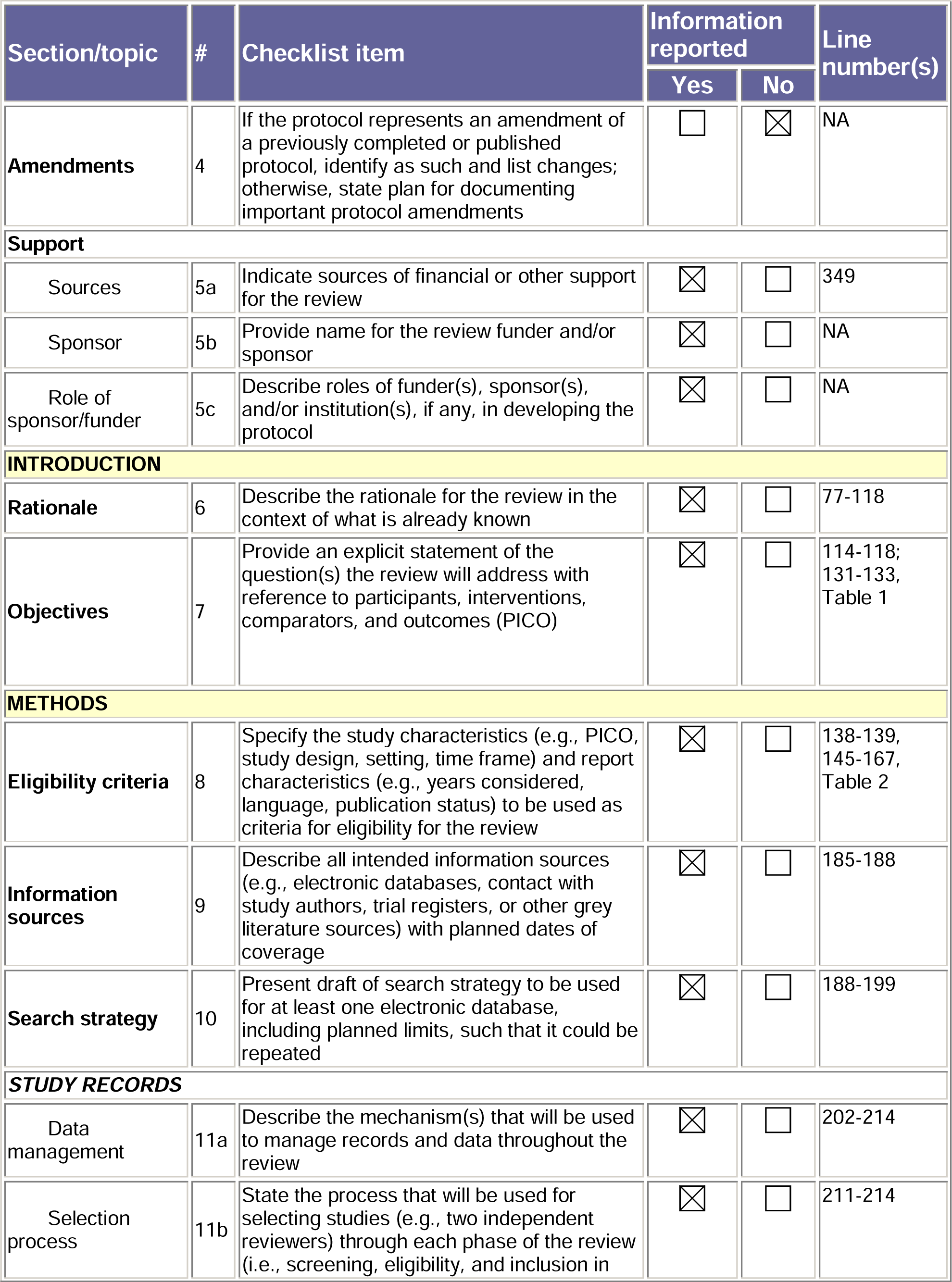
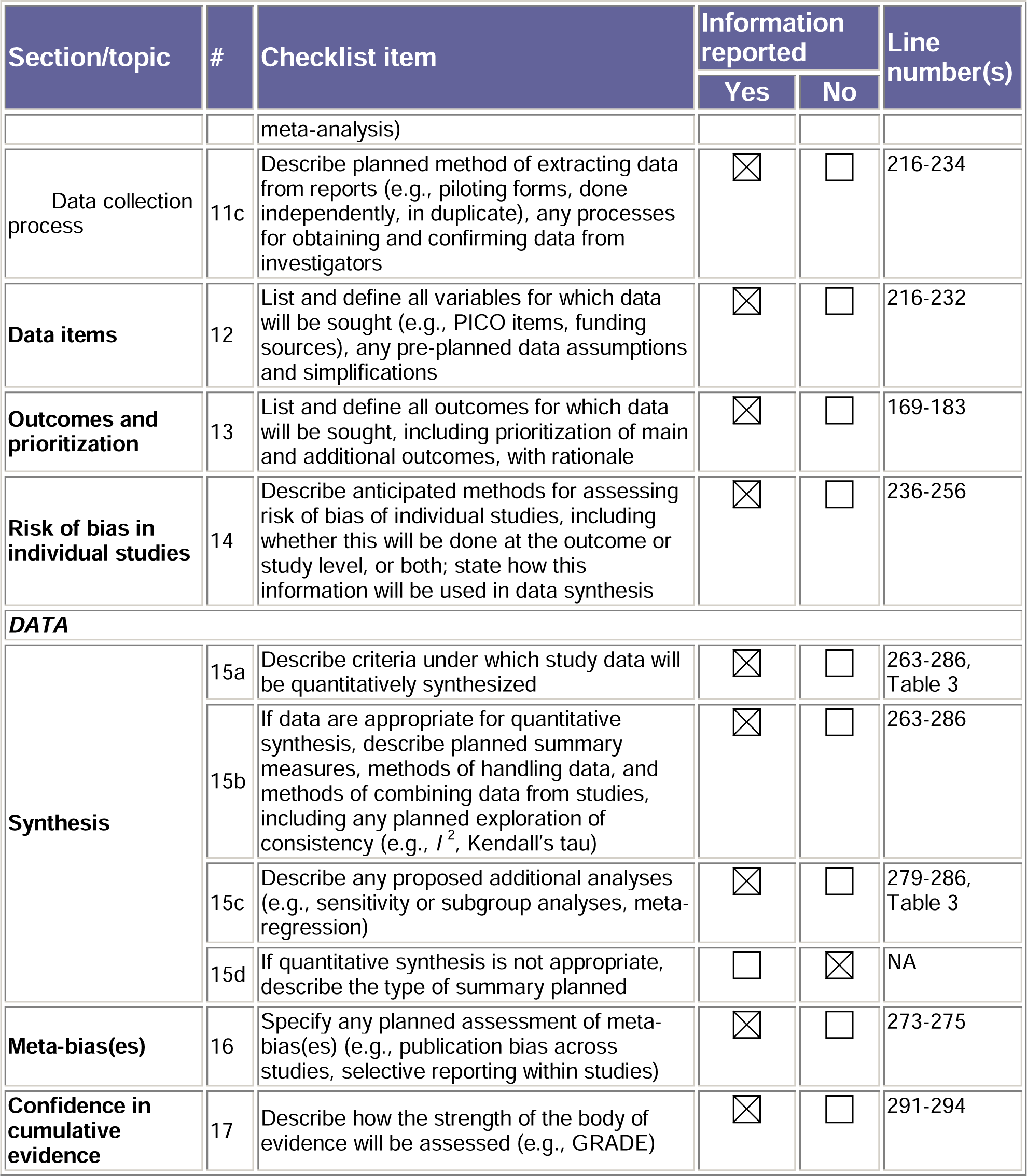

## ABBREVIATIONS

APACHE II: Acute Physiology and Chronic Health Evaluation-II
APT: Anti-platelet
ASA: Acetyl-salicylic acid
BiPAP: Bi-Level Positive Airway Pressure
CAP: Community Acquired Pneumonia
CI: Confidence Interval
COVID-19: Coronavirus Disease-19
CPAP: Continuous Positive Airway Pressure
CURB-65: Confusion, Urea, Respiratory Rate, Blood Pressure-65 score
DAPT: Dual Anti-platelet
ECLS: Extra-corporeal Life Support
GRADE: Grading of Recommendations, Assessment, Development, and Evaluations
HFNO: High Flow Nasal Oxygen
HR: Hazard Ratio
ICTRP: International Clinical Trials Registry Platform
ICU: Intensive Care Unit
IMV: Invasive Mechanical Ventilation
LOS: Length of Stay
LRTI: Lower Respiratory Tract Infection
MI: Myocardial Infarction
MOOSE: Meta-analysis of Observational Studies in Epidemiology
NIPPV: Non-Invasive Positive Pressure Ventilation
NOS: Newcastle Ottawa Scale
OR: Odds Ratio
PICO: Population Intervention Comparators and Outcomes
PRISMA: Preferred Reporting items for Systematic Reviews and Meta-analysis
PSI: Pneumonia Severity Index
RCT: Randomized Controlled Trial
RR: Risk Ratio
SOFA: Sequential Organ Failure Assessment
URTI: Upper Respiratory Tract Infection
VTE: Venous Thromboembolism
WMD: Weighted mean difference

